# Democracy, Inequality and Covid-19 Pandemic Outcomes: Age-standardised excess mortality and GDP growth in island and non-island jurisdictions

**DOI:** 10.64898/2026.01.22.26344652

**Authors:** Matt Boyd, Michael G Baker, Nick Wilson

## Abstract

**Background:** While democracy typically correlates with superior population health outcomes, and inequality adversely affects population wellbeing, their roles in pandemic performance remain contested, particularly across geographic context and when using methodologically robust metrics.

**Methods:** We examined associations between liberal democracy (V-Dem Liberal Democracy Index) and income inequality (Gini coefficient) with Covid-19 health and economic outcomes across 193 jurisdictions, stratified by island (n = 48) versus non-island (n = 145) status. Outcomes were age-standardised cumulative excess mortality (2020–2021) and GDP per capita growth (2019–2020, 2020–2021). Ordinary least squares regression models controlled for GDP per capita, population size, Global Health Security Index, and government corruption.

**Results:** Democracy predicted reduced excess mortality in islands (β = –5.92 ±2.20 SE, p = 0.013, adjusted R² = 0.37) but not non-islands (β = –0.47 ±0.65 SE, p = 0.47), confirmed by island interaction (β = –4.51 ±1.72, p = 0.0095). Higher inequality predicted increased mortality in non-islands (β = +0.052 ±0.019 SE, p = 0.009, adjusted R² = 0.50) and larger GDP contractions in 2019–2020 (β = –0.242 ±0.053 SE, p = 0.000013, adjusted R² = 0.22), but not in islands. Democracy showed no systematic association with economic trajectories.

**Conclusions:** Democracy’s pandemic benefits are geographically contingent, concentrated in island jurisdictions, while inequality’s adverse effects on health and economic outcomes are pervasive in non-island states. Preparedness strategies should account for these contextual dependencies to mitigate the impact of infectious disease and potential future global catastrophic biological risks.

**Key messages:**

- This research aimed to establish the relationship between jurisdiction regime type (level of democracy) as well as level of income inequality and Covid-19 pandemic health and economic outcomes.
- We found that a higher Gini inequality coefficient predicted greater cumulative excess mortality (2020–2021) and a larger initial economic contraction, while greater democracy predicted lower cumulative excess mortality in islands only.
- These findings suggest that non-specific factors such as inequality and democracy may drive pandemic outcomes and important policy relevant differences exist across jurisdiction types (island vs non-island).

## Introduction

The Covid-19 pandemic presented a global health crisis causing tens of millions of deaths. While preparedness factors such as the Global Health Security (GHS) Index ^[1]^, and strategic choices like border closures and an explicit exclusion/elimination strategy ^[2]^, influenced outcomes, non-specific factors including government corruption, social trust, and institutional quality also proved critical ^[2,3]^. Due to methodological issues, it remains unclear whether regime type, specifically the degree of democracy versus authoritarianism, or income inequality, predicted performance.

Democracy and lower inequality associate with better health across multiple indicators including mortality, life expectancy, and infant health ^[4–6]^. Democratisation through 1980–2016 drove 8-10% reductions in adult mortality relative to countries that remained autocratic ^[7]^. However, measuring Covid-19 outcomes presented unique methodological challenges that threatened to obscure genuine performance differences. Comparison of officially reported deaths with excess mortality across democracy levels, revealed large gaps in low-democracy regimes indicating unreported deaths but small gaps in high-democracy countries ^[8]^. Hence, studies using official Covid-19 statistics captured reporting behaviour rather than true mortality. Using excess mortality, which cannot be easily manipulated, democratic countries with higher government effectiveness reduced excess mortality ^[9]^.

Multiple studies using excess mortality and controlling for extensive confounders found consistent inverse associations between democracy quality and excess deaths, with effects ranging from 2.18 fewer deaths per 100,000 per democracy index point to strong effects for political culture dimensions ^[10–12]^.

While autocracies imposed more stringent lockdowns, democracies achieved 13-34% higher compliance with less restrictive measures through social capital and voluntary cooperation rather than coercion ^[13]^. Democracy’s health-protective pathways appear to include accountability, trust, and civic engagement, operating even under pandemic stress.

Limited evidence exists regarding economic outcomes. Countries with higher democratic maturity tended to suffer greater GDP per capita losses early in the pandemic, possibly reflecting more transparent reporting or proactive public health responses that temporarily constrained economic activity ^[14]^. However, more democratic states also enacted substantially larger fiscal stimulus packages ^[15]^.

Limited high-quality studies suggest income inequality may adversely affect pandemic outcomes, though evidence remains mixed. A systematic review found only two high-quality multi-country studies (n>100) examining the Gini coefficient and mortality ^[16]^. One study (n = 125) found a positive correlation between higher inequality and increased mortality ^[17]^, the other (n = 207) found higher inequality predicted lower mortality ^[18]^. However, this latter study used reported Covid-19 deaths, not excess mortality, and the former used an arbitrary metric of cumulative deaths. Other studies had methodological limitations including lack of age-standardisation ^[19,20]^.

A high-quality study of 29 European countries found the Gini coefficient was associated with increased excess mortality even when controlling for GDP, health expenditure, and vaccination rates ^[21]^. Another study examining 34 countries found “more vulnerable” countries (Gini >0.35) experienced more excess deaths per million than less vulnerable countries ^[22]^. Divergent regional and global findings may reflect genuine geographic heterogeneity or important methodological variations. Therefore, more analysis is needed.

Finally, recent research indicates that islands and non-islands experienced Covid-19 differently, with preparedness being important for non-islands while border restrictions proved more important for islands ^[1,2,23]^. Whether democracy and inequality’s effects on pandemic outcomes vary systematically between islands and non-islands remains unexplored.

## Aims

We aimed to determine the association between democracy and inequality as independent variables, and Covid-19-related health and macroeconomic outcomes as dependent variables (Figure 1). We evaluated democracy and income inequality in terms of island versus non-island contexts, which previous work showed to be an important distinction. To overcome gaps and methodological weaknesses of the existing literature we provided a global assessment and used robust age-standardised excess mortality data.

**Figure 1:**
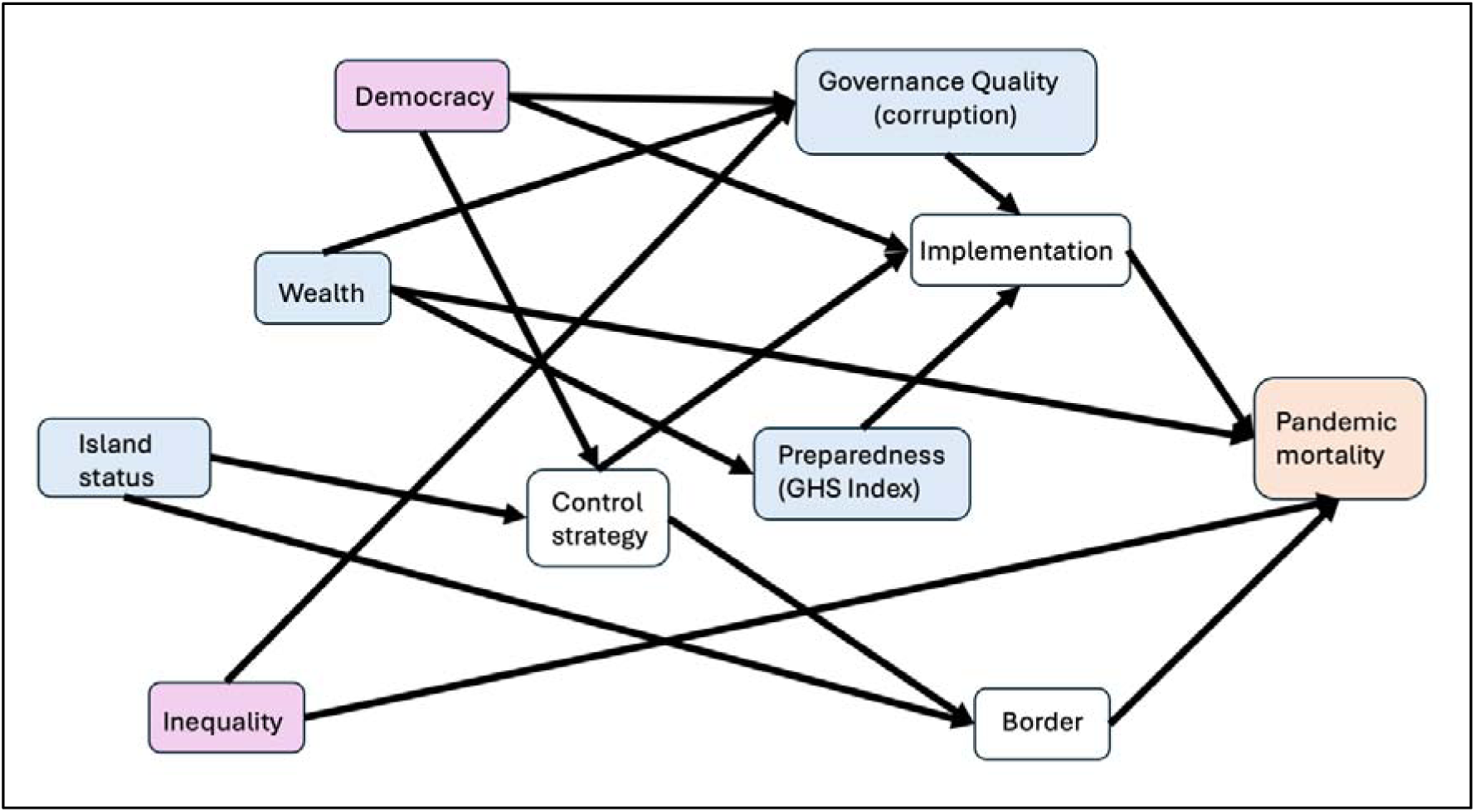
Causal assumptions used in the analysis (for outcome of pandemic-related excess mortality, shaded orange). Showing independent variables (red), covariates (blue), and mediating pathways (unshaded)

## Methods

### Jurisdictions studied

Our dataset comprised 193 sovereign jurisdictions categorised according to island (n = 48) and non-island status (n = 145). See Supplement for definition of islands and additional methodological details.

### Outcome variables

#### Excess mortality

Age-standardised cumulative excess mortality (2020 and 2021 combined), for all the jurisdictions was obtained from the GBD Study Demographics Collaborators ^[24]^. This age-standardised excess mortality variable represents the difference between observed all-cause deaths and expected deaths (based on pre-pandemic trends) during 2020–2021.

#### GDP per capita growth

We used growth in gross domestic product (GDP) (adjusted for purchasing power parity [PPP]) per capita to represent macroeconomic outcomes. GDP PPP per capita data for all jurisdictions was sourced from the World Bank’s World Development Indicators. Growth was measured as annual percentage change in five-year geometric mean of GDP PPP per capita.

### Independent variables

#### Democracy level

Following Kim ^[12]^, we used the V-Dem Liberal Democracy Index (LibDem Index). This dataset is collated and published by V-Dem and we used the pre-Covid-19 2019 iteration of data.

#### Income inequality

We used the Gini coefficient which is a measure of income inequality on a scale from 0 to 100, with higher values indicating higher inequality. Data for 2015–2019 were obtained from the World Bank, and the five-year mean used in analysis.

### Control variables

Covariates included in our models were: mean GDP per capita (as above) calculated for the five-year period ending in 2019; jurisdiction population size for 2019 obtained from the World Bank; GHS Index scores for 2019 ^[25]^ representing pandemic preparedness and previously correlated with pandemic outcomes ^[1]^; government corruption scores from the Covid-19 NP Collaborators, previously demonstrated to predict outcomes ^[3]^. Figure 1 illustrates our *a priori* causal assumptions.

### Analyses

#### Correlations among variables

Multicollinearity was assessed using variance inflation factors (VIFs) and Pearson correlation analysis (n = 116 countries, listwise deletion) to guide control variable selection. Control variables with absolute correlations exceeding 0.8 with other variables were excluded from final models, omitting the corruption index from LibDem models only.

#### Regression analysis

Ordinary least squares regression examined associations between LibDem (political) and Gini (economic) predictors and outcomes of excess mortality (signed cube-root transformed: sign·|value|^1/3) and GDP growth, with GDP per capita and population entered as natural logarithms. Six analysis families (inequality and mortality across islands, non-islands, all jurisdictions) were fitted first as unadjusted models (main predictor only), then as adjusted models adding log GDP per capita, log population, GHS index, and corruption index where applicable.

#### Interaction analysis

Interaction analyses tested whether associations between democracy (LibDem) or inequality (Gini 2015-2019) and outcomes (pandemic-related excess mortality, GDP growth 2019-20 and 2020-21) differed between island and non-island states by adding a binary island indicator and interaction term to fully-adjusted models. Models used heteroskedasticity-robust (HC3) standard errors to test whether slopes differed by island status conditional on controls.

## Results

### Descriptive data

Table 1 presents descriptive data across the three categories of jurisdiction.

**Table 1:** Descriptive data across jurisdiction categories.

|  | All Jurisdictions |  | Non-Islands |  | Islands |  |
| --- | --- | --- | --- | --- | --- | --- |
| Variable | Mean | N | Mean | N | Mean | N |
|  | (SD) |  | (SD) |  | (SD) |  |
|  | [Range] |  | [Range] |  | [Range] |  |
| Age-standardised EM (2020-2021) | 162.11<br>(154.44)<br>[-59.63, 897.42] | 193 | 194.33<br>(158.90)<br>[-29.4, 897.42] | 145 | 64.75<br>(84.79)<br>[-59.63, 333.68] | 48 |
| GDP growth 2019-2020 (%) | +0.39%<br>(2.6%)<br>[-9.2%, +10.0%] | 182 | +0.46%<br>(2.6%)<br>[-9.2%, +10.0%] | 137 | +0.18%<br>(2.6%)<br>[-5.8%, +5.6%] | 45 |
| GDP growth 2020-2021 (%) | +0.81%<br>(2.6%)<br>[-7.5%, +13.2%] | 182 | +0.99%<br>(2.6%)<br>[-7.5%, +13.2%] | 137 | +0.27%<br>(2.7%)<br>[-5.6%, +8.9%] | 45 |
| LibDem Index | 0.41<br>(0.26)<br>[0.01, 0.89] | 170 | 0.39<br>(0.26)<br>[0.01, 0.89] | 140 | 0.50<br>(0.24)<br>[0.05, 0.84] | 30 |
| Gini coefficient | 37.0<br>(7.20)<br>[24.6, 59.1] | 132 | 37.4<br>(7.55)<br>[24.6, 59.1] | 107 | 35.2<br>(5.20)<br>[26.7, 43.8] | 25 |
| Population | 40.7 million | 189 | 48.7 million | 143 | 15.8 million | 46 |
|  | (149 million) |  | (168 million) |  | (46.7 million) |  |
|  | [10,600, 1410 |  | [34,700, 1410 |  | [10,600, 272 million] |  |
|  | million] |  | million] |  |  |  |
| GHS Index | 40.5 | 191 | 42.2 | 144 | 35.25 | 47 |
| 2019 | (14.4) |  | (14.19) |  | (14.00) |  |
|  | [16.2, 83.5] |  | [16.2, 83.5] |  | [17.7, 77.9] |  |
| Government | -0.027 | 163 | 0.144 | 133 | -0.79 | 30 |
| Corruption | (1.39) |  | (1.35) |  | (1.33) |  |
| Index | [-2.79, 2.36] |  | [-2.79, 2.36] |  | [-2.76, 1.92] |  |

Note: EM = excess mortality per 100,000 population; N = sample size; SD = standard deviation; Range = [minimum, maximum]; GDP growth: growth in five-year mean GDP per capita year on year; GHS Index: Global Health Security Index score 2019; Government Corruption Index: perception of government corruption index as derived by the Covid-NP Collaborators (see Methods); LibDem Index: V-Dem Liberal Democracy Index.

### Regression analyses

#### Democracy (LibDem Index) and Covid-19 excess mortality

Full regression results across all models and variables can be found in the Supplement, Table S2. In the unadjusted analysis across all jurisdictions (n = 170), higher LibDem Index scores were strongly associated with lower age-standardised cumulative excess mortality (β = –4.25 ± 0.54 SE, p < 0.000001), see Table 2. The fully-adjusted model (n = 163) explained a moderate proportion of variance (adjusted R² = 0.39) with GDP per capita a strong negative predictor of mortality (β = –0.66, p = 0.0002). However, stratifying by island status revealed that among islands (n = 29), democracy exhibited a substantially larger negative association with mortality (β = –5.92 ± 2.20 SE, p = 0.013), see Figure 2. Whereas among non-islands (n = 134), there was little effect of democracy (β = –0.47 ± 0.65 SE, p = 0.47).

**Figure 2:**
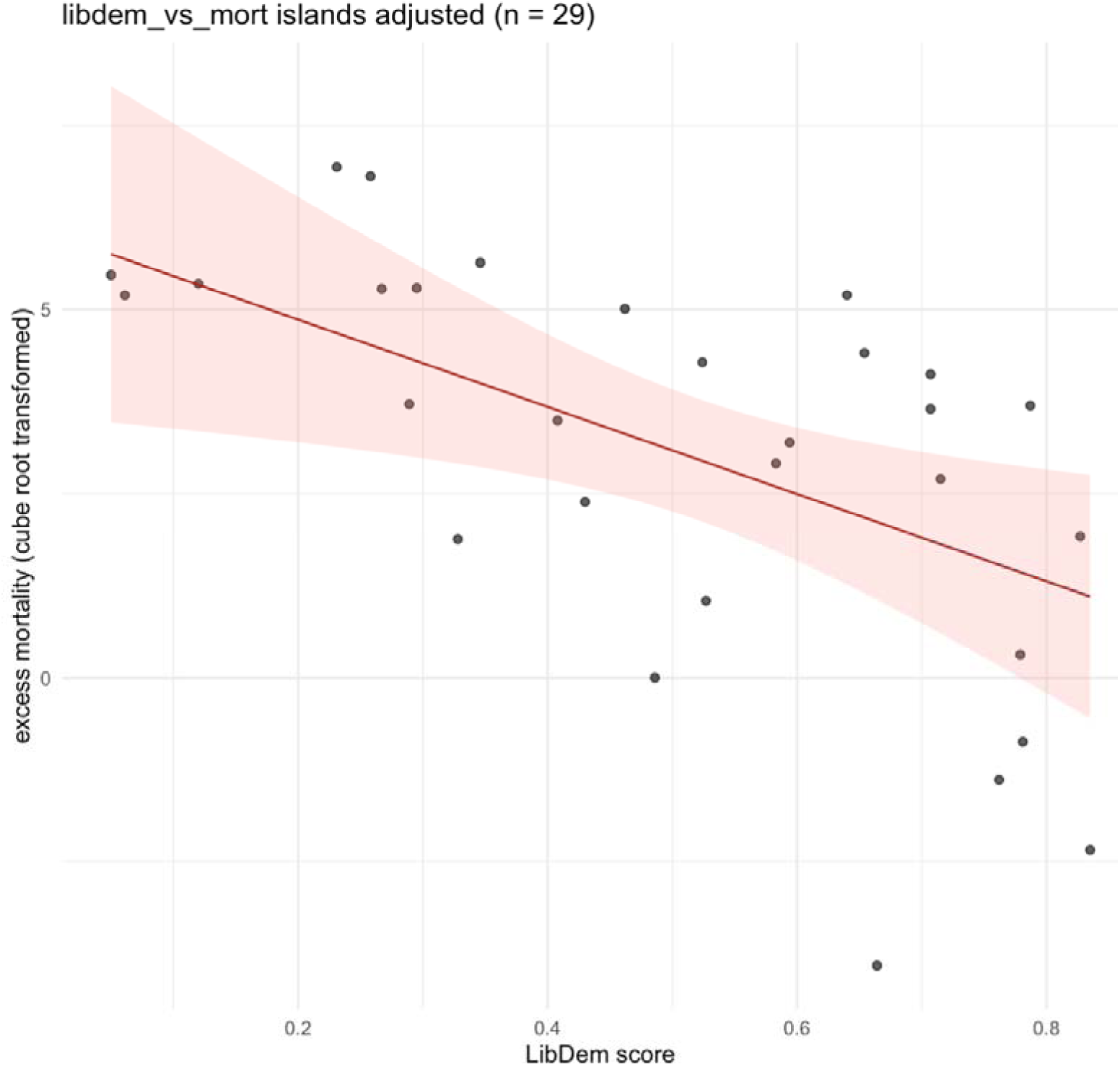
Relationship between LibDem score and cube root transformed cumulative excess mortality per 100,000 population (2020–2021) for island jurisdictions (fully-adjusted model controlling for GDP, GHS Index, and population size) [Alt text: Figure depicting the relationship between LibDem score and age-standardised cumulative excess mortality for island jurisdictions, showing excess mortality decreasing with higher LibDem scores]

**Table 2:**
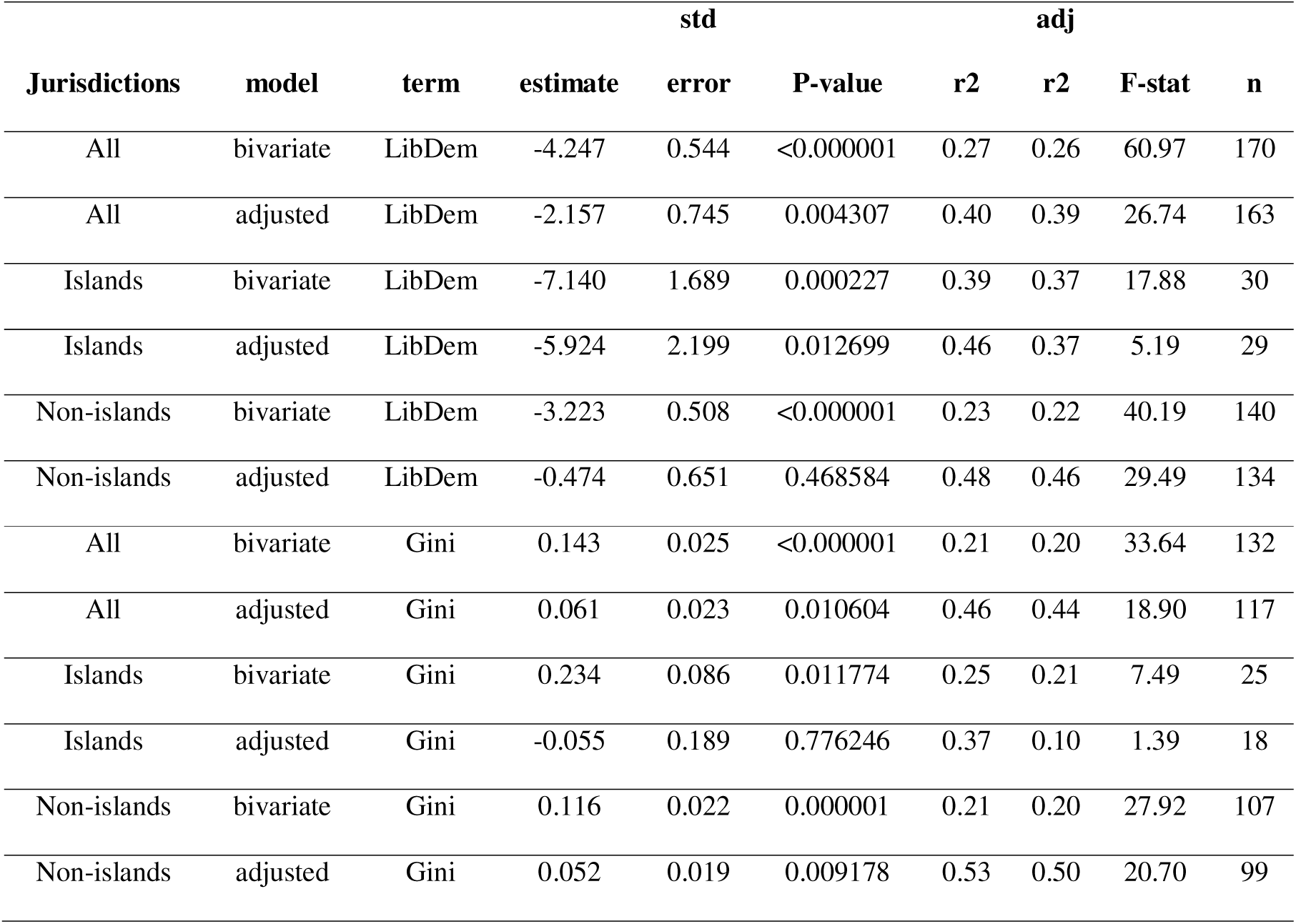
Regression results for the excess mortality models (LibDem and Gini, separately, vs age-standardised cumulative excess mortality 2020–2021, in fully-adjusted models controlling for GDP per capita, GHS Index, population size, and, in the Gini model, government corruption)

### Inequality (Gini) and Covid-19 excess mortality

Higher income inequality (Gini coefficient) was associated with higher age-standardised cumulative excess mortality in the fully-adjusted model (β = +0.0608 ± 0.0234 SE, t = 2.60, p = 0.0106, adj. R² = 0.44). GDP per capita (β = –0.569, p = 0.0128), and government corruption (β = +0.487, p = 0.0145) also showed associations.

This inequality–mortality relationship was absent among island jurisdictions (n = 18), (β = – 0.055, p = 0.78). However, among non-island jurisdictions (n = 99), inequality remained a predictor of excess mortality (β = +0.0518 ± 0.0195 SE, p = 0.0092, adj. R² = 0.50), see Figure 3, with predicted excess mortality decreasing by 15.8/100,000 (95% CI: -4.3 to -26.3) for a hypothetical excess mortality of 100/100,000 and a five-point Gini decrease (Supplementary Figure S2). GDP per capita (β = –0.431, p = 0.025) and GHS Index (β = – 0.036, p = 0.040) were negative predictors.

**Figure 3:**
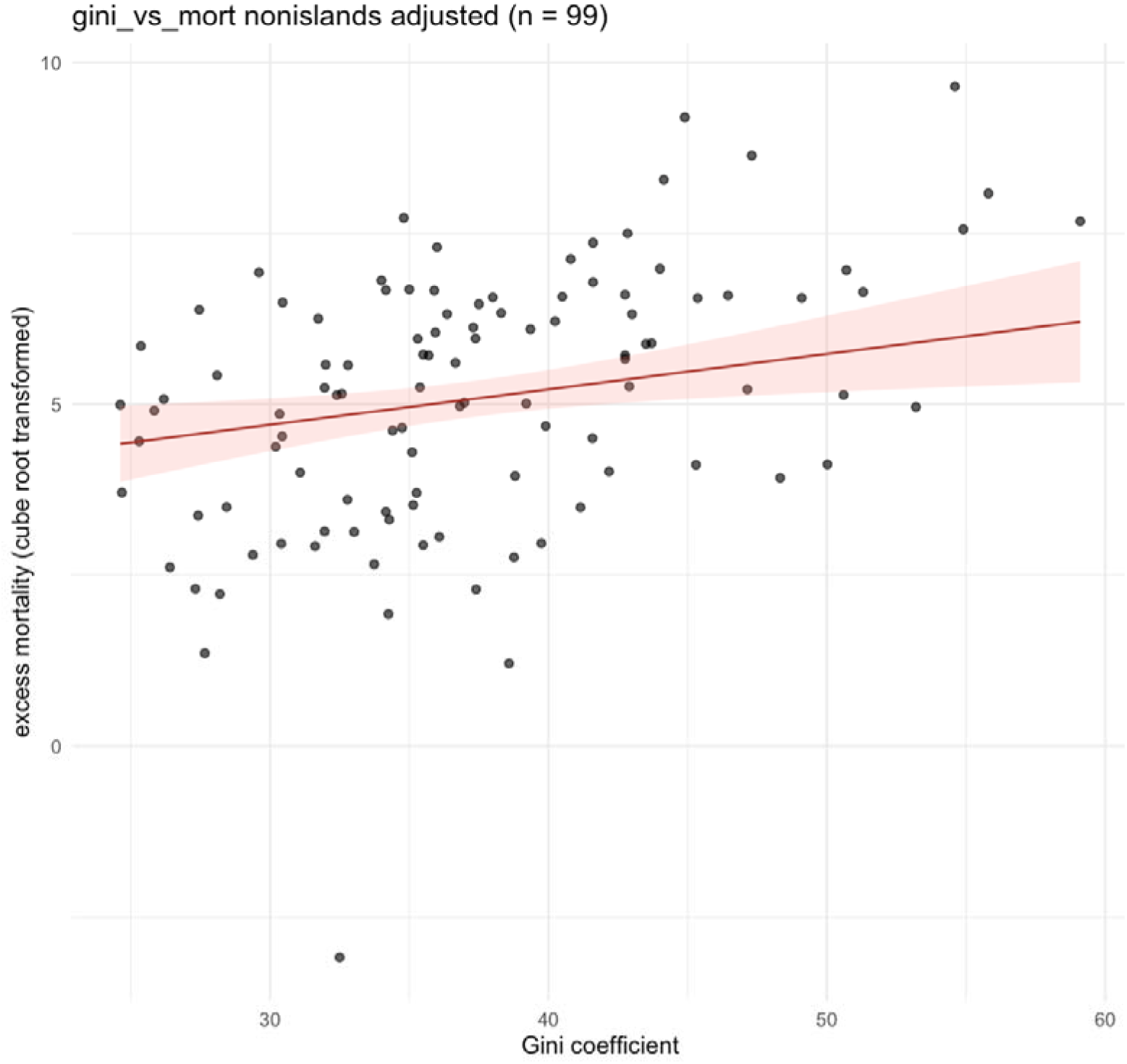
Relationship between Gini coefficient and cube root transformed cumulative excess mortality per 100,000 population (2020–2021) for non-island jurisdictions (fully adjusted model controlling for GDP, GHS Index, population size, and government corruption) [Alt text: Figure depicting the relationship between Gini coefficient and age-standardised cumulative excess mortality for non-island jurisdictions, showing excess mortality increasing with higher Gini coefficient]

### Democracy and GDP growth

Across all jurisdictions (n = 163), LibDem Index scores were not associated with GDP per-capita growth in the fully-adjusted model in either year under study, with the model explaining little variance. This pattern held for both islands and non-islands.

### Income inequality and GDP growth

Across all jurisdictions (n = 117), higher inequality was associated with larger GDP contractions in 2019–2020. In the adjusted model, Gini had a robust negative effect (β = – 0.225 ± 0.070 SE, p = 0.0018; adj. R² = 0.22). Population size was the only associated covariate (β = +1.44, p = 0.000077). A marginal positive association emerged for 2020–2021 GDP growth rebound (β = +0.137 ± 0.072 SE, p = 0.059), with higher pre-pandemic GDP predicting faster recovery (β = +2.13, p = 0.0026) and larger populations showing weaker growth in year two (β = –0.87, p = 0.017). This positive association only appeared after adjusting for structural controls.

In island jurisdictions (n = 18), inequality did not predict GDP change in either period (2019–2020: p = 0.339; 2020–2021: p = 0.517), with poor model fit in the second year (adj. R² = – 0.051).

Among non-islands (n = 99), inequality strongly predicted GDP decline in 2019–2020 (β = – 0.242 ± 0.053 SE, p = 0.000013, adj. R² = 0.22), see Figure 4, but was positively associated with 2020–2021 rebound (β = +0.146 ± 0.0583 SE, p = 0.014, adj. R² = 0.179), see Figure 5. Higher GDP per capita also predicted stronger rebound (p = 0.0028).

**Figure 4:**
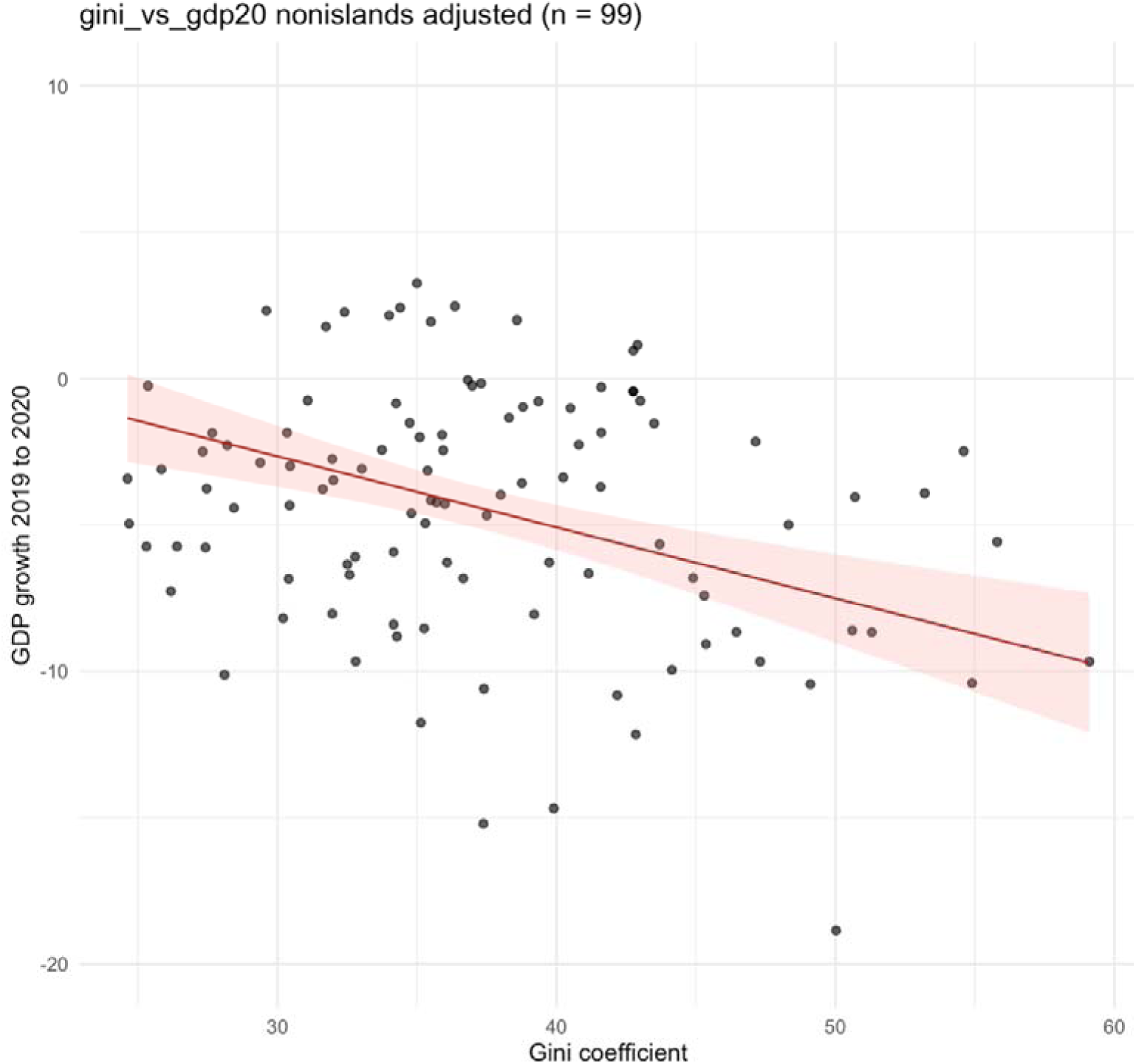
Relationship between Gini coefficient and GDP per capita growth 2019 to 2020 for non-island jurisdictions (fully adjusted model controlling for GDP, GHS Index, population, and government corruption) [Alt text: Figure depicting the relationship between Gini coefficient and GDP per capita growth 2019 to 2020 in non-island jurisdictions, showing GDP growth decreasing with higher Gini coefficient]

**Figure 5:**
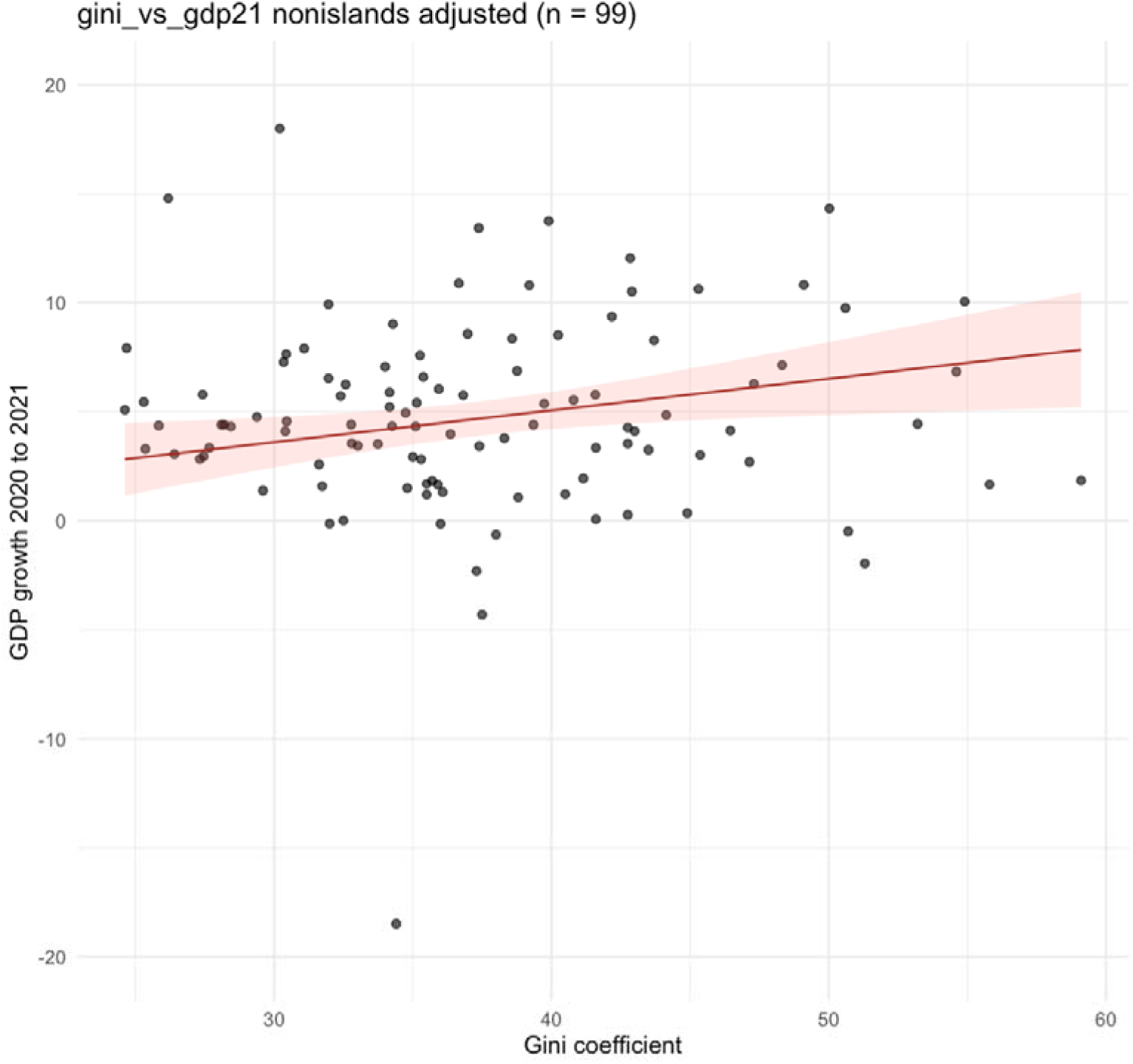
Relationship between Gini coefficient and GDP per capita growth 2021 to 2022 for non-island jurisdictions (fully-adjusted model controlling for GDP, GHS Index, population size, and government corruption) [Alt text: Figure depicting the relationship between Gini coefficient and GDP per capita growth 2020 to 20201in non-island jurisdictions, showing GDP growth increasing with higher Gini coefficient]

These results indicate inequality magnified Covid-19’s economic shock in 2020, primarily in non-island jurisdictions. More unequal non-island economies showed modestly stronger rebound after deeper initial contractions.

### Island interaction

In fully-adjusted models, island status moderated democracy’s effect on excess mortality (n = 163; LibDem*island: β = –4.51 ± 1.72, p = 0.0095). Democracy showed no main effect for non-islands (p = 0.22), but islands experienced markedly lower excess mortality at higher democratic scores. Among covariates, higher GDP per capita (p = 0.00012) and GHS capacity (p = 0.0248) predicted lower mortality. This suggests democracy’s protective effect was concentrated in islands, where border management and public-health autonomy may have heightened its impact. Although higher Gini positively associated with mortality (β = +0.053 ± 0.018, p = 0.0031), there was no Gini*island interaction (p = 0.432). Neither democracy nor inequality showed interactions with island status in predicting pandemic-year economic contraction or rebound.

## Discussion

Three main findings emerge from our results. First, liberal democracy was associated with lower excess mortality only in islands, formalised in a LibDem*island interaction. Second, higher income inequality was consistently associated with worse outcomes in non-islands with higher cumulative excess mortality 2020–21 and deeper GDP contractions in 2019–20, with modest rebound in 2020–21. Third, there was no robust evidence that democracy systematically shaped GDP trajectories once pre-pandemic structural factors were controlled for. Democracy’s pandemic benefits appear highly context-dependent, while inequality’s risks are pervasive, particularly in non-island states.

### Democracy, islands, and pandemic mortality

Democracy appears to “work differently” for islands, with a strong distinctive interaction between democracy and island status for Covid-19 mortality. This pattern suggests democratic governance effectiveness during pandemics depends on the interplay between political institutions and geographic constraints.

Democratic islands combining political legitimacy, cohesive social structures, and enforceable borders may have translated trust and accountability into timely interventions (stringent border controls, targeted quarantine, and clear public communication) through voluntary compliance without authoritarian coercion. This confluence of democracy and geography was most evident in the form of an explicit exclusion/elimination strategy which was associated with a negative excess mortality for the 2020-21 period in five countries, four of which were democratic islands ^[2]^.

The broader Covid-19 literature generally finds democracy protective for excess mortality when pooling all countries ^[9,11]^. Our analysis partially aligns but suggests this benefit is geographically contingent, concentrated in islands when applying age-standardisation, appropriate variable transformations, and robust controls (ie, GDP, population size, corruption, GHS Index).

### Inequality, mortality, and economic trajectories

Inequality’s role was more uniform, at least in non-island settings. Higher Gini coefficients were associated with higher excess mortality globally and among non-islands, with adjusted models explaining roughly half the variance. This result extends prior regional findings to a nearly global sample using age-standardised excess mortality.

The economic results reveal a similar structure. Inequality strongly predicted GDP decline from 2019 to 2020, yet more unequal non-island economies showed somewhat stronger rebound in 2020–21 once controls were included. This finding suggests an “overshoot” dynamic: high-inequality countries were more vulnerable to initial shocks but rebounded more strongly macroeconomically in year two, perhaps via aggressive reopening or concentrated sectoral recovery.

Importantly, this overshoot doesn’t mitigate underlying macroeconomic harm. Deeper initial contraction combined with higher mortality and stronger rebound is not equivalent to a gentler, more equal trajectory. Rather, inequality appears to act as a risk amplifier, exacerbating crisis vulnerability while fostering volatile recoveries that leave structural disparities intact or worsened. This underscores that redistributive and social protection policies are central to pandemic resilience, not peripheral.

### Democracy, economic performance, and the “lives vs livelihoods” narrative

One striking negative result is the absence of a robust relationship between democracy and macroeconomic outcomes. Once pre-pandemic GDP, population size, and health preparedness were controlled for, democracy did not predict either the 2019–20 contraction depth or 2020–21 recovery pace. Bivariate associations disappeared with structural controls, indicating confounding by wealth and baseline characteristics. This finding complicates narratives praising democracies for economic protection or authoritarian regimes for prioritising growth over health. Pre-existing economic structure, particularly baseline GDP per capita, emerges as the dominant recovery driver.

However, our results should not be over-interpreted as implying regime-type neutrality on all economic dimensions. We examine only aggregate GDP per-capita growth. Other outcomes, distributional effects, sector-specific impacts, employment, debt sustainability, or fiscal support composition, may differ systematically by regime type, as suggested by work showing larger fiscal packages in democracies ^[15]^. Detailed cross-national analysis of these dimensions remains largely absent and represents an important research agenda.

### Measurement timing, data quality, and methodological contributions

This study reinforces the critical importance of measurement choices in evaluating pandemic performance. Early studies using officially reported cases and deaths often showed authoritarian advantage, patterns that inverted when reliable excess mortality data became available ^[8]^. Our use of age-standardised cumulative excess mortality for 2020–21, with signed cube-root transformation to handle skew and negative values, reduces bias and estimation instability. Log transformations of GDP and population size, plus inclusion of the GHS Index and a corruption index, approximate a plausible structural model without overfitting.

This design inevitably involves trade-offs. Each covariate reduces sample size for complete-case analysis, particularly in inequality models where corruption data are less complete, raising generalisability and potential selection bias concerns. Excess mortality dataset choice matters as different global estimates vary in coverage, age-adjustment, and uncertainty handling. Our results therefore complement, rather than replace, earlier analyses using alternative sources and time windows. Full reconciliation of why some studies find strong global democracy effects while ours finds them primarily in islands requires side-by-side dataset comparison, standardised age-adjustment, and harmonised specifications.

Methodologically, our study combines (i) global coverage, (ii) age-standardised cumulative excess mortality, (iii) appropriate data transformations, (iv) theoretically grounded controls and causal diagram, (v) both democracy and inequality as predictors, and (vi) explicit island stratification. No prior work jointly satisfies all these conditions, particularly linking health and macroeconomic outcomes within a single framework.

### Potential implications for democratic resilience, inequality, and global catastrophic risk

Viewed alongside pre-Covid evidence that democracy generally improves population health, our findings suggest a more conditional picture under pandemic stress, with pronounced democracy benefits in island jurisdictions. This pattern suggests democracy’s effectiveness as a resilience enhancer depends on complementary conditions: administrative capacity, geographic manageability, and cohesive social structures. Where these are present, as in many islands, democracy and geography combine to produce exceptionally low pandemic-associated mortality.

Inequality’s consistent role as a risk multiplier has broader implications for global catastrophic biological risks. Historical work on societal collapse emphasises extractive institutions, corruption, and socio-economic stratification in undermining resilience, while recent theorising on long-term global risks highlights inequality’s dangers for collective action and social stability ^[26,27]^. Our results offer contemporary empirical support for these concerns.

## Limitations and future directions

Several limitations should be noted. First, this observational analysis cannot rule out unmeasured confounding from historical, cultural, geographic, or institutional factors that may influence both governance characteristics and pandemic outcomes. Second, sample sizes are modest in fully-adjusted models, particularly for islands (n ≈ 18–29), warranting cautious interpretation despite the statistically robust democracy*island interaction. Third, we focus on 2020–21, before widespread vaccination and long-term adaptation; later pandemic waves, virus variants, and vaccination campaigns may have altered these relationships. Fourth, economic outcomes are limited to aggregate GDP per-capita growth. Finally, our models explain up to 50% of variance, and spatial relations among jurisdictions are not fully captured, indicating our causal assumptions are necessarily incomplete.

Future work should incorporate spatial analyses to capture contagion dynamics and regional clustering, distinguish specific institutional features beyond broad democracy scores (electoral quality, civil liberties, centralised versus decentralised governance), extend longitudinally through later years to evaluate effects of democratic backsliding, fiscal exhaustion, and vaccination strategies, and employ a wider range of outcome measures.

## Conclusion

Our study suggests democracy’s pandemic-related benefits are conditional, emerging most clearly when combined with the greater opportunities for strong control policies provided by island geography. Additionally, high inequality consistently undermines health and economic performance, especially in non-island states. For policymakers, democratic quality must be coupled with capable institutions and reduced inequality to yield robust resilience. Investments in democratic governance, anti-corruption, and inequality reduction are therefore mutually reinforcing components of preparedness for future pandemics and large-scale global shocks, not competing priorities.

## Ethics statement

Our analysis employed only publicly available jurisdiction-level data, and as such was exempt from institutional ethical review.

## Acknowledgments

We thank Prof Austin Schumacher from the GBD Collaboration for data sharing.

## Author contributions

MB and NW conceived the study. MB, NW, and MGB contributed to the study design. MB collated data and performed the analysis, MB, NW, MGB contributed to drafting the manuscript, with each contributing important intellectual content.

## Supplementary data

Supplementary data are available at *IJE* online

## Data availability

Data and code used in this study are available here: https://adaptresearchwriting.com/wp-content/uploads/2026/01/260122-covid-gini-democracy-data-code.zip

## Use of Artificial Intelligence (AI) tools

AI was used to help iterate and improve R code for the analysis and to assist in reducing word count from an initial overlength paper. All AI outputs were closely scrutinised and revised by the authors before use in this study.

