## Supplement for "Democracy, Inequality and Covid-19 Pandemic Outcomes: Age-standardised excess mortality and GDP growth in island and non-island jurisdictions"

**Additional methodological details**

*Definition of island jurisdictions*

Island jurisdictions were defined as those surrounded by water, while ignoring engineered connections to other land masses. The group of island jurisdictions included: individual sovereign islands, island archipelagos (eg, Indonesia, Philippines), island continents (ie, Australia), and where the jurisdiction had a land border with another jurisdiction on the same island (eg, Ireland, UK, Timor-Leste, Papua New Guinea, Brunei, Dominican Republic, Haiti, Cyprus). We excluded jurisdictions with mixed characteristics, but where the capital city was on a continental land mass (eg, Malaysia, Corsica [France], Sardinia and Sicily [Italy]). We excluded non-sovereign island states due to the ambiguous role local government played in pandemic-related decision-making (as opposed to the colonial power). This definition of islands was consistent with earlier work on Covid-19 outcomes (Boyd, Baker, Kvalsvig, & Wilson, 2025).

*Excess mortality data*

The estimation methods for the GBD’s excess mortality data 2020–21 have been described in detail previously (GBD 2021 Demographics Collaborators, 2024). Data is standardised across age groups to enable comparison between populations with different age structures. The GBD Collaborators employed six weighted excess mortality models across a decade of data prior to the Covid-19 pandemic to attenuate any risk of arbitrary or outlier baseline data. Excess mortality is the best available measure of the true mortality impact from the Covid-19 and is not subject to the high levels of under ascertainment seen with Covid-19 mortality surveillance.

*GDP per capita growth*

The World Bank defines GDP per capita as GDP divided by midyear population. Specifically, we obtained the World Bank GDP per capita purchasing power parity (PPP) (constant 2017 international $) data, which included adjustment for inflation, ie, the World Bank dataset “NY.GDP.PCAP.PP.KD”. We then calculated GDP growth from 2019 to 2020, and from 2020 to 2021, defined as the percentage change in five-year geometric means.

*Causal model and covariate selection*

We developed a causal diagram (Figure 1, main text) to make explicit our assumptions linking pre-pandemic political and social conditions to age-standardised cumulative excess mortality in 2020–2021. The diagram included liberal democracy (V-Dem Liberal Democracy Index, 2019) and income inequality (Gini coefficient; mean 2015–2019) as primary exposures, with pre-pandemic GDP per capita, population size, island status, preparedness capacity (Global Health Security Index, 2019), and governance quality (proxied by a corruption index) as additional nodes.

In the diagram, governance quality was conceptualised as an institutional channel through which democracy may influence implementation effectiveness of pandemic control measures; therefore, to estimate the total effect of democracy, we did not condition on corruption in democracy models (also avoiding collinearity given strong correlation between democracy and corruption).

For inequality, we estimated a controlled direct effect (not mediated by governance quality) by additionally conditioning on the corruption index, thereby blocking the pathway from inequality to mortality operating through governance/implementation.

Across models, we included pre-pandemic GDP per capita, population size, preparedness (GHS Index), and island status (and, where relevant, exposure x island interaction terms) to address confounding and to assess effect modification by island status. We conceptualised “control strategy” as being on one of the pathways for how more democratic jurisdictions might impact pandemic mortality (but we could not include this variable in modelling given that so few jurisdictions had explicit control strategies, see (Boyd et al., 2025)).

Conditioning on corruption in inequality models is intended as a robustness/mechanism test consistent with a causal mechanism in which governance quality mediates part of the relationship between inequality and mortality.

*Causal diagram nodes and relations*

\# Nodes

Island

Wealth

LibDem

Gini

GovQuality

GHSI

Strategy

Borders

Implementation

Mortality

\# Relations

Wealth -> GovQuality

Wealth -> GHSI

LibDem -> GovQuality

Gini -> GovQuality

Island -> Strategy

Island -> Borders

Strategy -> Borders

Strategy -> Implementation

LibDem -> Strategy

LibDem -> Implementation

GovQuality -> Implementation

GHSI -> Implementation

Wealth -> Mortality

Gini -> Mortality

Borders -> Mortality

Implementation -> Mortality

**Additional results**

*Correlations and collinearity*

Table S1 reports the Pearson correlations among the key variables. Based on our *a priori* Pearson correlation threshold (see Methods), the perceived government corruption index was excluded from the LibDem regressions due to its strong correlation with democracy (Pearson’s r = 0.797, which we took to be concerningly close to our 0.8 *a priori* threshold), but was retained in the Gini models.

**Table S1**: Pearson’s r correlations among variables

|  | **LibDem** | **Gini** | **Corruption** | **GHS_2019** | **log_GDP_2019** | **log_population** |
| --- | --- | --- | --- | --- | --- | --- |
| **LibDem** | - | -0.216 | -0.797 | 0.592 | 0.58 | -0.238 |
| **Gini** | -0.216 | - | 0.357 | -0.317 | -0.398 | 0.157 |
| **Corruption** | -0.797 | 0.357 | - | -0.633 | -0.734 | 0.234 |
| **GHS_2019** | 0.592 | -0.317 | -0.633 | - | 0.686 | 0.29 |
| **log_GDP_2019** | 0.58 | -0.398 | -0.734 | 0.686 | - | -0.119 |
| **log_population** | -0.238 | 0.157 | 0.234 | 0.29 | -0.119 | - |

For the LibDem models (with covariates log GDP per capita, log population size, and the GHS Index), VIF values ranged from approximately 1.7 to 3.5 (LibDem ≈ 2.0, log GDP ≈ 2.3, log population ≈ 1.7, GHS ≈ 3.5; *n* = 163), well below conventional concern thresholds. For the income inequality models (Gini coefficient plus log GDP, log population size, GHS Index, and corruption index), VIFs were likewise modest (≈1.2–3.3; *n* = 117).

**Personalist Autocracies and pandemic mortality**

Seven of the jurisdictions studied can be considered personalist autocracies (ie, authoritarian regimes under the control of a single autocrat whose personal authority outweighs institutions, laws, and elite constraints; and where decision-making is typically highly centralised, unpredictable, and closely tied to the ruler’s personal preferences and survival). These personalist autocracies had excess mortality of 229 per 100,000 population vs 160 per 100,000 for all other jurisdictions (p < 0.01), and versus 193 per 100,000 for non-island jurisdictions (p = 0.14).

**Additional figures**

**Figure S1**: Visualisation of relationships between LibDem score or Gini coefficient (independent variables) and age-standardised cumulative excess mortality 2020–2021 (“mort”), GDP per capita growth 2019–2020 (“gdp20”), and 2020–2021 (“gdp21”); for non-island jurisdictions only.

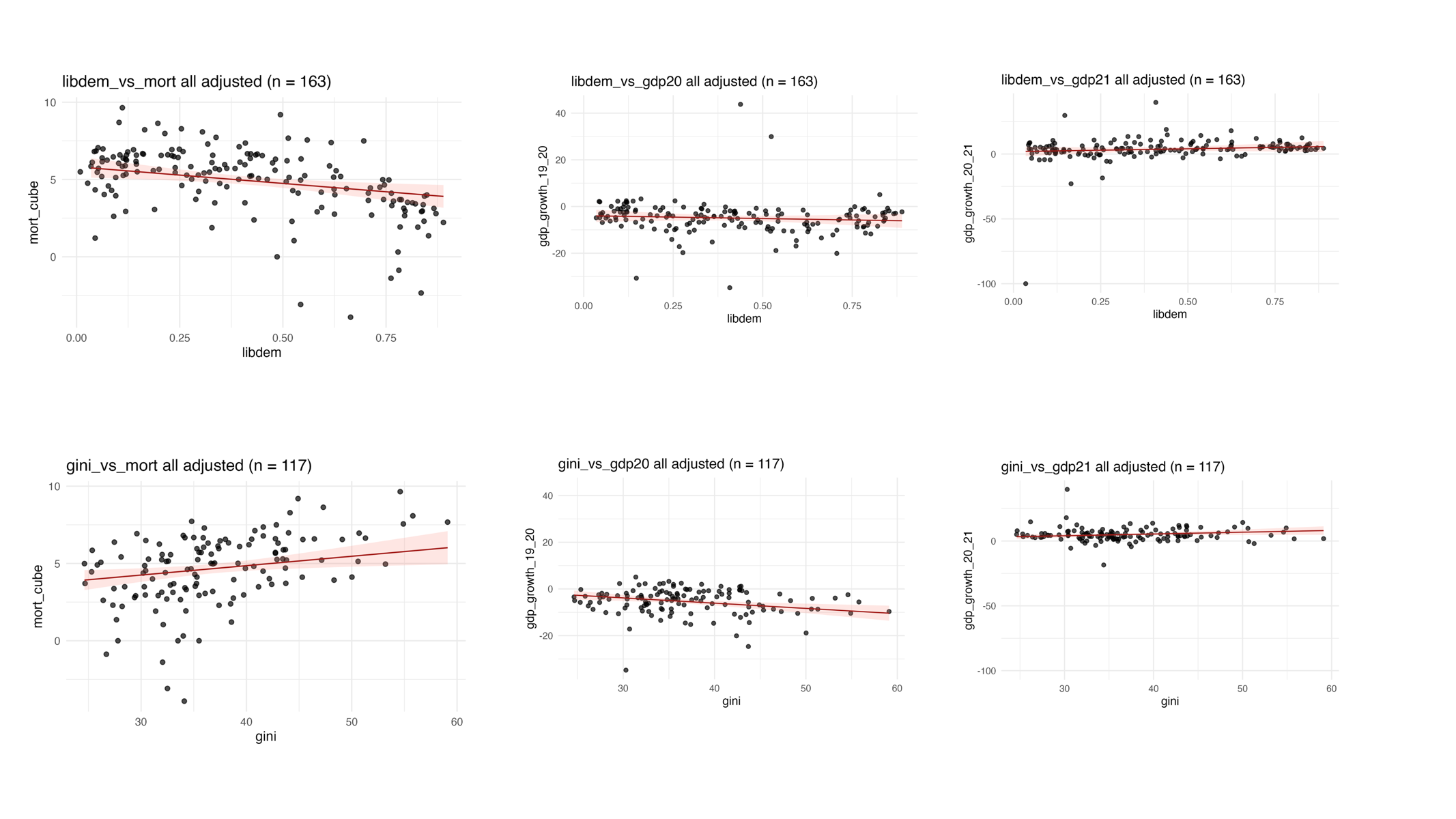

**Figure S2:** Predicted relationship between Gini coefficient (independent variable) and age-standardised cumulative excess mortality 2020–2021 for non-island jurisdictions only.

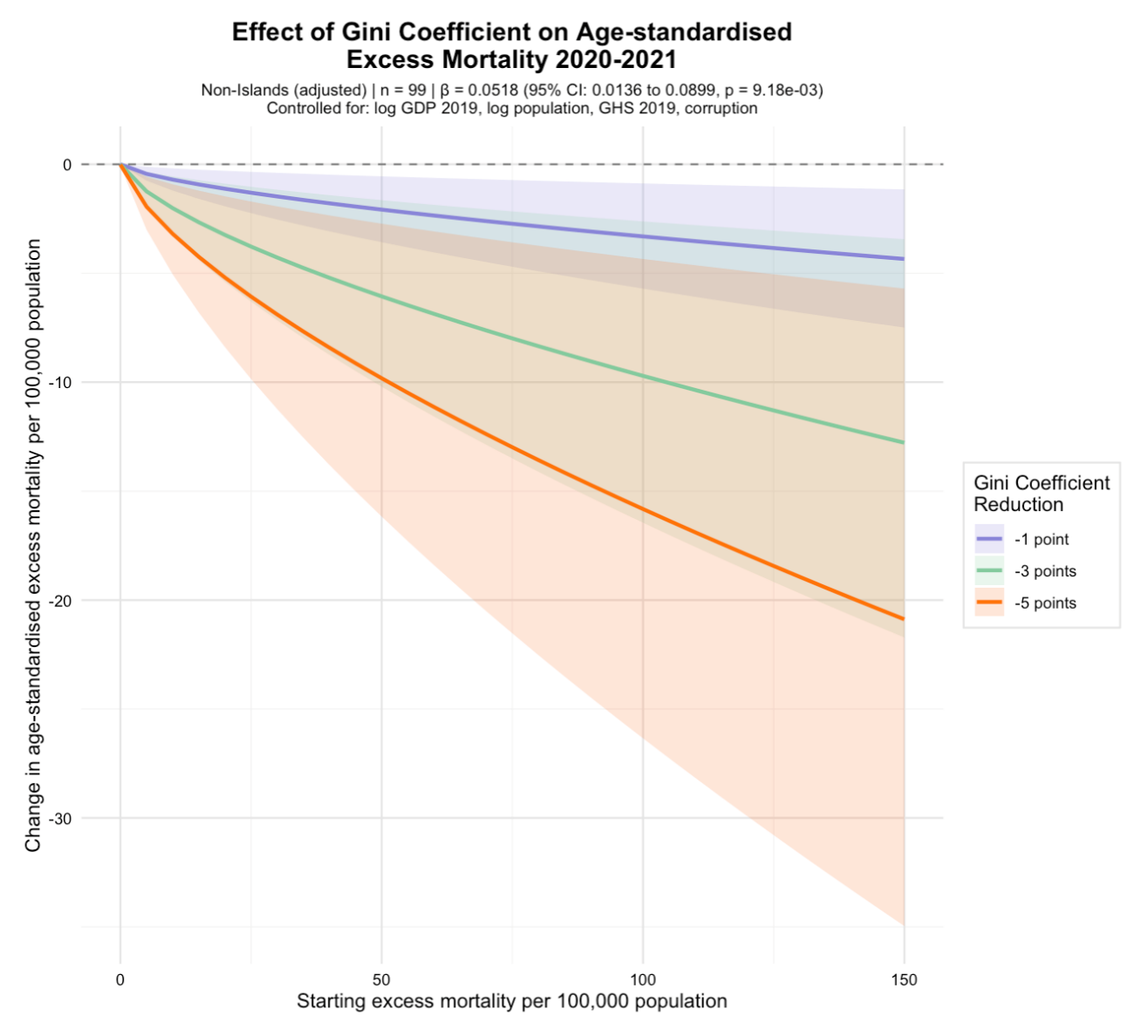

**Table S2**: Complete regression results across six aims (A1-A6)

| **analysis** | **stratum_model** | **term** | **estimate** | **std.error** | **t statistic** | **p.value** | **r2** | **Adj r2** | **f_stat** | **p_overall** | **n** |
| --- | --- | --- | --- | --- | --- | --- | --- | --- | --- | --- | --- |
| A1_libdem_mort | all_unadjusted | (Intercept) | 6.66927274 | 0.26339555 | 25.3203703 | 3.10E-59 | 0.27 | 0.26 | 60.97 | 5.95E-13 | 170 |
| A1_libdem_mort | all_unadjusted | libdem | -4.2474021 | 0.54396901 | -7.8081692 | 5.95E-13 | 0.27 | 0.26 | 60.97 | 5.95E-13 | 170 |
| A1_libdem_mort | all_adjusted | (Intercept) | 10.4628876 | 2.32001505 | 4.5098361 | 1.26E-05 | 0.40 | 0.39 | 26.74 | 6.00E-17 | 163 |
| A1_libdem_mort | all_adjusted | libdem | -2.1567906 | 0.74459171 | -2.8966084 | 0.00430747 | 0.40 | 0.39 | 26.74 | 6.00E-17 | 163 |
| A1_libdem_mort | all_adjusted | log_gdp_2019 | -0.6616933 | 0.1738988 | -3.8050481 | 2.02E-04 | 0.40 | 0.39 | 26.74 | 6.00E-17 | 163 |
| A1_libdem_mort | all_adjusted | log_pop | 0.13993939 | 0.10311954 | 1.3570598 | 0.1766977 | 0.40 | 0.39 | 26.74 | 6.00E-17 | 163 |
| A1_libdem_mort | all_adjusted | ghs_2019 | -0.0161122 | 0.01796039 | -0.8970964 | 0.37103237 | 0.40 | 0.39 | 26.74 | 6.00E-17 | 163 |
| A1_libdem_mort | islands_unadjusted | (Intercept) | 6.64428582 | 0.93730206 | 7.08873493 | 1.03E-07 | 0.39 | 0.37 | 17.88 | 2.27E-04 | 30 |
| A1_libdem_mort | islands_unadjusted | libdem | -7.1404562 | 1.6886435 | -4.2285161 | 2.27E-04 | 0.39 | 0.37 | 17.88 | 2.27E-04 | 30 |
| A1_libdem_mort | islands_adjusted | (Intercept) | 8.96926261 | 7.89592387 | 1.1359358 | 0.26719988 | 0.46 | 0.37 | 5.19 | 0.00369011 | 29 |
| A1_libdem_mort | islands_adjusted | libdem | -5.9235489 | 2.19932312 | -2.6933509 | 0.01269871 | 0.46 | 0.37 | 5.19 | 0.00369011 | 29 |
| A1_libdem_mort | islands_adjusted | log_gdp_2019 | -0.6326351 | 0.5929045 | -1.0670101 | 0.29658354 | 0.46 | 0.37 | 5.19 | 0.00369011 | 29 |
| A1_libdem_mort | islands_adjusted | log_pop | 0.21387519 | 0.34887457 | 0.61304324 | 0.54561396 | 0.46 | 0.37 | 5.19 | 0.00369011 | 29 |
| A1_libdem_mort | islands_adjusted | ghs_2019 | -4.45E-04 | 0.06467577 | -0.0068788 | 0.99456839 | 0.46 | 0.37 | 5.19 | 0.00369011 | 29 |
| A1_libdem_mort | nonislands_unadjusted | (Intercept) | 6.59085151 | 0.23772758 | 27.7243871 | 2.85E-58 | 0.23 | 0.22 | 40.19 | 3.05E-09 | 140 |
| A1_libdem_mort | nonislands_unadjusted | libdem | -3.2229676 | 0.50836297 | -6.3398946 | 3.05E-09 | 0.23 | 0.22 | 40.19 | 3.05E-09 | 140 |
| A1_libdem_mort | nonislands_adjusted | (Intercept) | 12.4545037 | 1.98888191 | 6.26206292 | 5.21E-09 | 0.48 | 0.46 | 29.49 | 2.04E-17 | 134 |
| A1_libdem_mort | nonislands_adjusted | libdem | -0.4735164 | 0.6513885 | -0.7269339 | 0.46858358 | 0.48 | 0.46 | 29.49 | 2.04E-17 | 134 |
| A1_libdem_mort | nonislands_adjusted | log_gdp_2019 | -0.5119684 | 0.14757268 | -3.4692624 | 7.10E-04 | 0.48 | 0.46 | 29.49 | 2.04E-17 | 134 |
| A1_libdem_mort | nonislands_adjusted | log_pop | -0.0059082 | 0.09037224 | -0.0653761 | 0.94797573 | 0.48 | 0.46 | 29.49 | 2.04E-17 | 134 |
| A1_libdem_mort | nonislands_adjusted | ghs_2019 | -0.0475499 | 0.01538414 | -3.0908406 | 0.00244585 | 0.48 | 0.46 | 29.49 | 2.04E-17 | 134 |
| A2_gini_mort | all_unadjusted | (Intercept) | -0.6567231 | 0.92705276 | -0.7083988 | 0.47996592 | 0.21 | 0.20 | 33.64 | 4.77E-08 | 132 |
| A2_gini_mort | all_unadjusted | gini | 0.14266679 | 0.02459686 | 5.80020444 | 4.77E-08 | 0.21 | 0.20 | 33.64 | 4.77E-08 | 132 |
| A2_gini_mort | all_adjusted | (Intercept) | 7.57051291 | 2.80440584 | 2.69950689 | 0.00803099 | 0.46 | 0.44 | 18.90 | 1.47E-13 | 117 |
| A2_gini_mort | all_adjusted | gini | 0.0607636 | 0.02337476 | 2.59953871 | 0.01060371 | 0.46 | 0.44 | 18.90 | 1.47E-13 | 117 |
| A2_gini_mort | all_adjusted | log_gdp_2019 | -0.5689734 | 0.22474287 | -2.5316641 | 0.0127517 | 0.46 | 0.44 | 18.90 | 1.47E-13 | 117 |
| A2_gini_mort | all_adjusted | log_pop | 0.0383101 | 0.11634817 | 0.32927124 | 0.74257117 | 0.46 | 0.44 | 18.90 | 1.47E-13 | 117 |
| A2_gini_mort | all_adjusted | ghs_2019 | -0.0034607 | 0.01936098 | -0.1787479 | 0.85846152 | 0.46 | 0.44 | 18.90 | 1.47E-13 | 117 |
| A2_gini_mort | all_adjusted | corruption | 0.48668723 | 0.19593413 | 2.48393287 | 0.01448803 | 0.46 | 0.44 | 18.90 | 1.47E-13 | 117 |
| A2_gini_mort | islands_unadjusted | (Intercept) | -5.7836824 | 3.04737607 | -1.8979221 | 0.07032775 | 0.25 | 0.21 | 7.49 | 0.01177375 | 25 |
| A2_gini_mort | islands_unadjusted | gini | 0.23430019 | 0.08563538 | 2.73602089 | 0.01177375 | 0.25 | 0.21 | 7.49 | 0.01177375 | 25 |
| A2_gini_mort | islands_adjusted | (Intercept) | 21.0090142 | 23.9516638 | 0.87714216 | 0.39763011 | 0.37 | 0.10 | 1.39 | 0.29665492 | 18 |
| A2_gini_mort | islands_adjusted | gini | -0.0549935 | 0.18917783 | -0.2906974 | 0.77624567 | 0.37 | 0.10 | 1.39 | 0.29665492 | 18 |
| A2_gini_mort | islands_adjusted | log_gdp_2019 | -1.4476992 | 1.73524349 | -0.8342917 | 0.42041504 | 0.37 | 0.10 | 1.39 | 0.29665492 | 18 |
| A2_gini_mort | islands_adjusted | log_pop | -0.2698176 | 0.52531932 | -0.513626 | 0.61683892 | 0.37 | 0.10 | 1.39 | 0.29665492 | 18 |
| A2_gini_mort | islands_adjusted | ghs_2019 | 0.07623842 | 0.10548862 | 0.72271703 | 0.48370191 | 0.37 | 0.10 | 1.39 | 0.29665492 | 18 |
| A2_gini_mort | islands_adjusted | corruption | 1.33565405 | 0.90589513 | 1.4744025 | 0.16611913 | 0.37 | 0.10 | 1.39 | 0.29665492 | 18 |
| A2_gini_mort | nonislands_unadjusted | (Intercept) | 0.80132593 | 0.83462952 | 0.96009776 | 0.33921168 | 0.21 | 0.20 | 27.92 | 6.89E-07 | 107 |
| A2_gini_mort | nonislands_unadjusted | gini | 0.11556203 | 0.02186906 | 5.28427065 | 6.89E-07 | 0.21 | 0.20 | 27.92 | 6.89E-07 | 107 |
| A2_gini_mort | nonislands_adjusted | (Intercept) | 9.90746915 | 2.42392464 | 4.08736682 | 9.25E-05 | 0.53 | 0.50 | 20.70 | 7.64E-14 | 99 |
| A2_gini_mort | nonislands_adjusted | gini | 0.05179719 | 0.01946532 | 2.6609993 | 0.00917762 | 0.53 | 0.50 | 20.70 | 7.64E-14 | 99 |
| A2_gini_mort | nonislands_adjusted | log_gdp_2019 | -0.4312912 | 0.18924283 | -2.2790359 | 0.02495263 | 0.53 | 0.50 | 20.70 | 7.64E-14 | 99 |
| A2_gini_mort | nonislands_adjusted | log_pop | -0.0587979 | 0.1048333 | -0.56087 | 0.57623498 | 0.53 | 0.50 | 20.70 | 7.64E-14 | 99 |
| A2_gini_mort | nonislands_adjusted | ghs_2019 | -0.0359212 | 0.01728456 | -2.0782223 | 0.04044379 | 0.53 | 0.50 | 20.70 | 7.64E-14 | 99 |
| A2_gini_mort | nonislands_adjusted | corruption | 0.19725911 | 0.17318437 | 1.13901217 | 0.25762349 | 0.53 | 0.50 | 20.70 | 7.64E-14 | 99 |
| A3_libdem_gdp20 | all_unadjusted | (Intercept) | -3.7471233 | 1.07399732 | -3.4889504 | 6.25E-04 | 0.01 | 0.00 | 1.67 | 0.19862548 | 164 |
| A3_libdem_gdp20 | all_unadjusted | libdem | -2.8124718 | 2.17892468 | -1.2907614 | 0.19862548 | 0.01 | 0.00 | 1.67 | 0.19862548 | 164 |
| A3_libdem_gdp20 | all_adjusted | (Intercept) | 1.9830728 | 9.62725118 | 0.20598536 | 0.83706752 | 0.04 | 0.02 | 1.86 | 0.12035345 | 163 |
| A3_libdem_gdp20 | all_adjusted | libdem | -2.4438134 | 3.08979523 | -0.7909306 | 0.43017016 | 0.04 | 0.02 | 1.86 | 0.12035345 | 163 |
| A3_libdem_gdp20 | all_adjusted | log_gdp_2019 | -1.3505901 | 0.7216192 | -1.8716106 | 0.06310907 | 0.04 | 0.02 | 1.86 | 0.12035345 | 163 |
| A3_libdem_gdp20 | all_adjusted | log_pop | 0.16023542 | 0.42791005 | 0.37446053 | 0.70856381 | 0.04 | 0.02 | 1.86 | 0.12035345 | 163 |
| A3_libdem_gdp20 | all_adjusted | ghs_2019 | 0.09751013 | 0.07452936 | 1.30834524 | 0.19265632 | 0.04 | 0.02 | 1.86 | 0.12035345 | 163 |
| A3_libdem_gdp20 | islands_unadjusted | (Intercept) | -8.1054951 | 4.87412624 | -1.6629637 | 0.10788671 | 0.00 | -0.03 | 0.06 | 0.81559547 | 29 |
| A3_libdem_gdp20 | islands_unadjusted | libdem | 2.03365601 | 8.63537068 | 0.23550304 | 0.81559547 | 0.00 | -0.03 | 0.06 | 0.81559547 | 29 |
| A3_libdem_gdp20 | islands_adjusted | (Intercept) | -4.0640782 | 38.9101426 | -0.1044478 | 0.91768199 | 0.06 | -0.10 | 0.36 | 0.83140371 | 29 |
| A3_libdem_gdp20 | islands_adjusted | libdem | 4.06568611 | 10.8379941 | 0.37513271 | 0.71085658 | 0.06 | -0.10 | 0.36 | 0.83140371 | 29 |
| A3_libdem_gdp20 | islands_adjusted | log_gdp_2019 | -1.7947464 | 2.92176051 | -0.6142688 | 0.54481735 | 0.06 | -0.10 | 0.36 | 0.83140371 | 29 |
| A3_libdem_gdp20 | islands_adjusted | log_pop | 0.58306751 | 1.719211 | 0.33914832 | 0.73744768 | 0.06 | -0.10 | 0.36 | 0.83140371 | 29 |
| A3_libdem_gdp20 | islands_adjusted | ghs_2019 | 0.08595784 | 0.31871423 | 0.26970193 | 0.78969497 | 0.06 | -0.10 | 0.36 | 0.83140371 | 29 |
| A3_libdem_gdp20 | nonislands_unadjusted | (Intercept) | -3.3291829 | 0.9992174 | -3.3317903 | 0.00111745 | 0.01 | 0.01 | 1.87 | 0.17405592 | 135 |
| A3_libdem_gdp20 | nonislands_unadjusted | libdem | -2.8679498 | 2.09859921 | -1.366602 | 0.17405592 | 0.01 | 0.01 | 1.87 | 0.17405592 | 135 |
| A3_libdem_gdp20 | nonislands_adjusted | (Intercept) | 5.52722029 | 9.44500626 | 0.58520028 | 0.55943532 | 0.04 | 0.01 | 1.17 | 0.3257577 | 134 |
| A3_libdem_gdp20 | nonislands_adjusted | libdem | -3.1567978 | 3.09338045 | -1.020501 | 0.30940095 | 0.04 | 0.01 | 1.17 | 0.3257577 | 134 |
| A3_libdem_gdp20 | nonislands_adjusted | log_gdp_2019 | -1.159382 | 0.70080829 | -1.6543497 | 0.10048769 | 0.04 | 0.01 | 1.17 | 0.3257577 | 134 |
| A3_libdem_gdp20 | nonislands_adjusted | log_pop | -0.1032447 | 0.42916898 | -0.2405689 | 0.81027119 | 0.04 | 0.01 | 1.17 | 0.3257577 | 134 |
| A3_libdem_gdp20 | nonislands_adjusted | ghs_2019 | 0.08641381 | 0.07305776 | 1.18281489 | 0.23905792 | 0.04 | 0.01 | 1.17 | 0.3257577 | 134 |
| A4_gini_gdp20 | all_unadjusted | (Intercept) | -1.0313678 | 2.57612387 | -0.4003564 | 0.68955624 | 0.02 | 0.01 | 2.93 | 0.08942012 | 131 |
| A4_gini_gdp20 | all_unadjusted | gini | -0.1171339 | 0.06844539 | -1.7113487 | 0.08942012 | 0.02 | 0.01 | 2.93 | 0.08942012 | 131 |
| A4_gini_gdp20 | all_adjusted | (Intercept) | -11.332381 | 8.42918394 | -1.344422 | 0.18155302 | 0.25 | 0.22 | 7.50 | 4.24E-06 | 117 |
| A4_gini_gdp20 | all_adjusted | gini | -0.2246759 | 0.07025737 | -3.1978983 | 0.0018041 | 0.25 | 0.22 | 7.50 | 4.24E-06 | 117 |
| A4_gini_gdp20 | all_adjusted | log_gdp_2019 | -0.9389451 | 0.67550814 | -1.3899833 | 0.1673152 | 0.25 | 0.22 | 7.50 | 4.24E-06 | 117 |
| A4_gini_gdp20 | all_adjusted | log_pop | 1.43661894 | 0.3497069 | 4.10806571 | 7.66E-05 | 0.25 | 0.22 | 7.50 | 4.24E-06 | 117 |
| A4_gini_gdp20 | all_adjusted | ghs_2019 | -0.0037617 | 0.05819317 | -0.064642 | 0.94857534 | 0.25 | 0.22 | 7.50 | 4.24E-06 | 117 |
| A4_gini_gdp20 | all_adjusted | corruption | -0.1036144 | 0.58891791 | -0.1759402 | 0.86066148 | 0.25 | 0.22 | 7.50 | 4.24E-06 | 117 |
| A4_gini_gdp20 | islands_unadjusted | (Intercept) | -0.3441951 | 12.4431863 | -0.0276613 | 0.97817088 | 0.02 | -0.02 | 0.52 | 0.47954045 | 25 |
| A4_gini_gdp20 | islands_unadjusted | gini | -0.2513218 | 0.34967032 | -0.7187393 | 0.47954045 | 0.02 | -0.02 | 0.52 | 0.47954045 | 25 |
| A4_gini_gdp20 | islands_adjusted | (Intercept) | -195.38413 | 72.2497656 | -2.7042874 | 0.01915682 | 0.56 | 0.37 | 3.04 | 0.05331655 | 18 |
| A4_gini_gdp20 | islands_adjusted | gini | 0.56824576 | 0.57065155 | 0.99578414 | 0.33901129 | 0.56 | 0.37 | 3.04 | 0.05331655 | 18 |
| A4_gini_gdp20 | islands_adjusted | log_gdp_2019 | 12.4565243 | 5.23433098 | 2.37977391 | 0.03477712 | 0.56 | 0.37 | 3.04 | 0.05331655 | 18 |
| A4_gini_gdp20 | islands_adjusted | log_pop | 3.85984875 | 1.58461633 | 2.43582543 | 0.03139651 | 0.56 | 0.37 | 3.04 | 0.05331655 | 18 |
| A4_gini_gdp20 | islands_adjusted | ghs_2019 | -0.4704987 | 0.31820454 | -1.4786045 | 0.16500962 | 0.56 | 0.37 | 3.04 | 0.05331655 | 18 |
| A4_gini_gdp20 | islands_adjusted | corruption | -2.0961693 | 2.73261647 | -0.7670924 | 0.45785386 | 0.56 | 0.37 | 3.04 | 0.05331655 | 18 |
| A4_gini_gdp20 | nonislands_unadjusted | (Intercept) | 0.72149137 | 2.00710062 | 0.35946946 | 0.71997278 | 0.06 | 0.05 | 6.92 | 0.0098185 | 106 |
| A4_gini_gdp20 | nonislands_unadjusted | gini | -0.1385586 | 0.05267213 | -2.6305865 | 0.0098185 | 0.06 | 0.05 | 6.92 | 0.0098185 | 106 |
| A4_gini_gdp20 | nonislands_adjusted | (Intercept) | 1.05932416 | 6.55480934 | 0.16161022 | 0.87196339 | 0.26 | 0.22 | 6.66 | 2.41E-05 | 99 |
| A4_gini_gdp20 | nonislands_adjusted | gini | -0.242495 | 0.05263837 | -4.6068113 | 1.30E-05 | 0.26 | 0.22 | 6.66 | 2.41E-05 | 99 |
| A4_gini_gdp20 | nonislands_adjusted | log_gdp_2019 | -0.893207 | 0.51175297 | -1.7453871 | 0.08422036 | 0.26 | 0.22 | 6.66 | 2.41E-05 | 99 |
| A4_gini_gdp20 | nonislands_adjusted | log_pop | 0.95073333 | 0.28349161 | 3.35365592 | 0.00115509 | 0.26 | 0.22 | 6.66 | 2.41E-05 | 99 |
| A4_gini_gdp20 | nonislands_adjusted | ghs_2019 | -0.0816904 | 0.04674115 | -1.7477197 | 0.08381312 | 0.26 | 0.22 | 6.66 | 2.41E-05 | 99 |
| A4_gini_gdp20 | nonislands_adjusted | corruption | -0.6678714 | 0.46832749 | -1.4260778 | 0.15719409 | 0.26 | 0.22 | 6.66 | 2.41E-05 | 99 |
| A5_libdem_gdp21 | all_unadjusted | (Intercept) | 0.33917216 | 1.50044778 | 0.22604729 | 0.82144958 | 0.04 | 0.04 | 6.94 | 0.00923371 | 164 |
| A5_libdem_gdp21 | all_unadjusted | libdem | 8.02084685 | 3.04410694 | 2.63487683 | 0.00923371 | 0.04 | 0.04 | 6.94 | 0.00923371 | 164 |
| A5_libdem_gdp21 | all_adjusted | (Intercept) | -4.7137176 | 13.6131095 | -0.3462631 | 0.72960566 | 0.05 | 0.03 | 2.24 | 0.06756072 | 163 |
| A5_libdem_gdp21 | all_adjusted | libdem | 4.37371997 | 4.36902705 | 1.00107414 | 0.31832164 | 0.05 | 0.03 | 2.24 | 0.06756072 | 163 |
| A5_libdem_gdp21 | all_adjusted | log_gdp_2019 | 0.87348918 | 1.02038276 | 0.85604071 | 0.39327163 | 0.05 | 0.03 | 2.24 | 0.06756072 | 163 |
| A5_libdem_gdp21 | all_adjusted | log_pop | -0.2012301 | 0.60507264 | -0.3325717 | 0.73989841 | 0.05 | 0.03 | 2.24 | 0.06756072 | 163 |
| A5_libdem_gdp21 | all_adjusted | ghs_2019 | 0.03853437 | 0.10538587 | 0.36565025 | 0.71511514 | 0.05 | 0.03 | 2.24 | 0.06756072 | 163 |
| A5_libdem_gdp21 | islands_unadjusted | (Intercept) | 3.9167058 | 3.95494186 | 0.99033208 | 0.3308045 | 0.00 | -0.04 | 0.04 | 0.8416989 | 29 |
| A5_libdem_gdp21 | islands_unadjusted | libdem | 1.41296359 | 7.00687412 | 0.20165391 | 0.8416989 | 0.00 | -0.04 | 0.04 | 0.8416989 | 29 |
| A5_libdem_gdp21 | islands_adjusted | (Intercept) | -7.2009881 | 29.7050679 | -0.2424161 | 0.81051735 | 0.17 | 0.03 | 1.19 | 0.34223372 | 29 |
| A5_libdem_gdp21 | islands_adjusted | libdem | -6.9692279 | 8.27402133 | -0.8423024 | 0.4079344 | 0.17 | 0.03 | 1.19 | 0.34223372 | 29 |
| A5_libdem_gdp21 | islands_adjusted | log_gdp_2019 | 2.37368938 | 2.23055195 | 1.06417131 | 0.29784131 | 0.17 | 0.03 | 1.19 | 0.34223372 | 29 |
| A5_libdem_gdp21 | islands_adjusted | log_pop | -0.7669838 | 1.31249274 | -0.5843719 | 0.56442342 | 0.17 | 0.03 | 1.19 | 0.34223372 | 29 |
| A5_libdem_gdp21 | islands_adjusted | ghs_2019 | 0.09934129 | 0.24331517 | 0.40828235 | 0.6866862 | 0.17 | 0.03 | 1.19 | 0.34223372 | 29 |
| A5_libdem_gdp21 | nonislands_unadjusted | (Intercept) | -0.0847277 | 1.63979674 | -0.0516696 | 0.95886946 | 0.05 | 0.04 | 6.83 | 0.00997852 | 135 |
| A5_libdem_gdp21 | nonislands_unadjusted | libdem | 9.0027952 | 3.44397138 | 2.61407375 | 0.00997852 | 0.05 | 0.04 | 6.83 | 0.00997852 | 135 |
| A5_libdem_gdp21 | nonislands_adjusted | (Intercept) | -4.7021843 | 15.6557527 | -0.3003487 | 0.76439471 | 0.05 | 0.02 | 1.78 | 0.13624921 | 134 |
| A5_libdem_gdp21 | nonislands_adjusted | libdem | 7.00522194 | 5.12749256 | 1.36620812 | 0.17425053 | 0.05 | 0.02 | 1.78 | 0.13624921 | 134 |
| A5_libdem_gdp21 | nonislands_adjusted | log_gdp_2019 | 0.53747496 | 1.16163833 | 0.46268701 | 0.64436826 | 0.05 | 0.02 | 1.78 | 0.13624921 | 134 |
| A5_libdem_gdp21 | nonislands_adjusted | log_pop | -0.0245886 | 0.71137733 | -0.0345647 | 0.97248025 | 0.05 | 0.02 | 1.78 | 0.13624921 | 134 |
| A5_libdem_gdp21 | nonislands_adjusted | ghs_2019 | 0.01871387 | 0.12109831 | 0.15453451 | 0.87742982 | 0.05 | 0.02 | 1.78 | 0.13624921 | 134 |
| A6_gini_gdp21 | all_unadjusted | (Intercept) | 4.86014438 | 2.49582165 | 1.94731237 | 0.05366989 | 0.00 | -0.01 | 0.00 | 0.97408604 | 131 |
| A6_gini_gdp21 | all_unadjusted | gini | 0.00215826 | 0.06631183 | 0.03254713 | 0.97408604 | 0.00 | -0.01 | 0.00 | 0.97408604 | 131 |
| A6_gini_gdp21 | all_adjusted | (Intercept) | -8.4653578 | 8.60723379 | -0.9835167 | 0.32749214 | 0.18 | 0.14 | 4.80 | 5.21E-04 | 117 |
| A6_gini_gdp21 | all_adjusted | gini | 0.13697825 | 0.07174141 | 1.90933292 | 0.05880082 | 0.18 | 0.14 | 4.80 | 5.21E-04 | 117 |
| A6_gini_gdp21 | all_adjusted | log_gdp_2019 | 2.12669454 | 0.68977692 | 3.08316282 | 0.00258422 | 0.18 | 0.14 | 4.80 | 5.21E-04 | 117 |
| A6_gini_gdp21 | all_adjusted | log_pop | -0.8685559 | 0.35709377 | -2.4322907 | 0.0166019 | 0.18 | 0.14 | 4.80 | 5.21E-04 | 117 |
| A6_gini_gdp21 | all_adjusted | ghs_2019 | 0.05617327 | 0.05942239 | 0.94532172 | 0.34654789 | 0.18 | 0.14 | 4.80 | 5.21E-04 | 117 |
| A6_gini_gdp21 | all_adjusted | corruption | 1.09334392 | 0.60135764 | 1.81812595 | 0.0717417 | 0.18 | 0.14 | 4.80 | 5.21E-04 | 117 |
| A6_gini_gdp21 | islands_unadjusted | (Intercept) | 9.35004568 | 12.029464 | 0.77726203 | 0.44492569 | 0.00 | -0.04 | 0.10 | 0.75016276 | 25 |
| A6_gini_gdp21 | islands_unadjusted | gini | -0.1089399 | 0.33804417 | -0.3222652 | 0.75016276 | 0.00 | -0.04 | 0.10 | 0.75016276 | 25 |
| A6_gini_gdp21 | islands_adjusted | (Intercept) | 57.7737068 | 96.0285579 | 0.60163047 | 0.55861536 | 0.26 | -0.05 | 0.84 | 0.54934338 | 18 |
| A6_gini_gdp21 | islands_adjusted | gini | -0.506051 | 0.75846398 | -0.667205 | 0.51726611 | 0.26 | -0.05 | 0.84 | 0.54934338 | 18 |
| A6_gini_gdp21 | islands_adjusted | log_gdp_2019 | 0.05034741 | 6.95705033 | 0.00723689 | 0.99434477 | 0.26 | -0.05 | 0.84 | 0.54934338 | 18 |
| A6_gini_gdp21 | islands_adjusted | log_pop | -3.0315113 | 2.10614414 | -1.4393655 | 0.17561873 | 0.26 | -0.05 | 0.84 | 0.54934338 | 18 |
| A6_gini_gdp21 | islands_adjusted | ghs_2019 | 0.42671967 | 0.42293179 | 1.00895624 | 0.33290786 | 0.26 | -0.05 | 0.84 | 0.54934338 | 18 |
| A6_gini_gdp21 | islands_adjusted | corruption | 6.50049582 | 3.63197328 | 1.78979726 | 0.09873209 | 0.26 | -0.05 | 0.84 | 0.54934338 | 18 |
| A6_gini_gdp21 | nonislands_unadjusted | (Intercept) | 4.08345172 | 2.20984224 | 1.84784762 | 0.06746623 | 0.00 | -0.01 | 0.11 | 0.7398716 | 106 |
| A6_gini_gdp21 | nonislands_unadjusted | gini | 0.0193064 | 0.05799266 | 0.33291104 | 0.7398716 | 0.00 | -0.01 | 0.11 | 0.7398716 | 106 |
| A6_gini_gdp21 | nonislands_adjusted | (Intercept) | -10.541647 | 7.25500676 | -1.4530168 | 0.14958505 | 0.22 | 0.18 | 5.28 | 2.59E-04 | 99 |
| A6_gini_gdp21 | nonislands_adjusted | gini | 0.14584256 | 0.0582613 | 2.50324939 | 0.01405008 | 0.22 | 0.18 | 5.28 | 2.59E-04 | 99 |
| A6_gini_gdp21 | nonislands_adjusted | log_gdp_2019 | 1.74147686 | 0.56641942 | 3.07453596 | 0.0027675 | 0.22 | 0.18 | 5.28 | 2.59E-04 | 99 |
| A6_gini_gdp21 | nonislands_adjusted | log_pop | -0.5510004 | 0.31377474 | -1.7560381 | 0.08237405 | 0.22 | 0.18 | 5.28 | 2.59E-04 | 99 |
| A6_gini_gdp21 | nonislands_adjusted | ghs_2019 | 0.05364812 | 0.05173413 | 1.03699674 | 0.30242618 | 0.22 | 0.18 | 5.28 | 2.59E-04 | 99 |
| A6_gini_gdp21 | nonislands_adjusted | corruption | 0.64728212 | 0.51835514 | 1.24872325 | 0.21490056 | 0.22 | 0.18 | 5.28 | 2.59E-04 | 99 |

**Table notes**: adjr2: adjusted r-squared metric; corruption: government corruption index score; f_stat: F-statistic; gdp20: growth in gross domestic product (PPP) per capita 2019 to 2020; gdp21: growth in gross domestic product (PPP) per capita 2020 to 2021; ghs_2019: Global Health Security Index score (2019); gini: Gini income inequality coefficient; mort: age-standardised cumulative excess mortality 2020–2021; pop: population size; r2: r-squared metric.
